# Effects of *MAPT* rs17649553 on Structural Network Integrity and Verbal Memory in Parkinson’s Disease

**DOI:** 10.1101/2023.01.14.23284559

**Authors:** Zhichun Chen, Bin Wu, Guanglu Li, Liche Zhou, Lina Zhang, Jun Liu

## Abstract

**Background:** Currently, over 90 genetic loci have been found to be associated with Parkinson’s disease (PD) in genome-wide association studies, nevertheless, the effects of these genetic variants on the clinical features and brain structure of PD patients are largely unknown.

**Objective:** This study investigated the effects of microtubule-associated protein tau (*MAPT*), rs17649553 (C>T), a genetic variant associated with reduced PD risk, on the functional and structural networks of PD patients.

**Methods:** Totally 83 PD subjects from Parkinson’s Progression Markers Initiative database were included for this study. They all received structural and functional magnetic resonance imaging and whole exome sequencing. The effects of *MAPT* rs17649553 on brain structural and functional networks were systematically assessed.

**Results:** *MAPT* rs17649553 T allele was associated with better verbal memory in PD patients. In addition, *MAPT* rs17649553 significantly reshaped the topology of gray matter covariance network and white matter network but not that of functional network. Both the network metrics in gray matter covariance network and white matter network were correlated with verbal memory, however, the mediation analysis showed that it was the small-worldness topology in white matter network that mediated the effects of *MAPT* rs17649553 on verbal memory.

**Conclusion:** In sum, we proposed that *MAPT* rs17649553 T allele was associated with superior structural network topology and better verbal memory in PD. Future studies are needed to determine the role of *MAPT* rs17649553 in PD initiation and progression.

## INTRODUCTION

Functional magnetic resonance imaging (fMRI) is an excellent tool to assess and monitor how the brain changes during the onset and progression of neuropsychiatric diseases, including bipolar disorder, schizophrenia, Parkinson’s disease (PD), and Alzheimer’s disease (Matsuo *et al*., 2012; Skudlarski *et al*., 2013; Montembeault *et al*., 2016; Nestor *et al*., 2017; Imperiale *et al*., 2018; Kamagata *et al*., 2018; Buckner and DiNicola, 2019; Filippi *et al*., 2020). Previously, three types of brain networks have been reported in neuroimaging studies, including functional network, white matter network, and gray matter covariance network (Bullmore and Sporns, 2009; Tijms *et al*., 2012; Alexander-Bloch *et al*., 2013; Filippi *et al*., 2013; Montembeault *et al*., 2016). These networks provide an integral measurement of brain organization and function. Due to different computation methodologies, functional network and structural network are not usually consistent (Lawrence *et al*., 2018; Luo *et al*., 2020), but they provide complementary information about the changes of brain structure and function (Bullmore and Sporns, 2009). Importantly, recent studies have combined the functional and structural networks to identify neuroimaging alterations related to clinical characteristics of diseases (Fishman *et al*., 2015; Hong *et al*., 2017; Imperiale *et al*., 2018; Filippi *et al*., 2020). To understand the genetic basis of human brain, neuroimaging genetics approaches have been utilized to reveal the associations between molecular genetics and brain structure or function (Satizabal *et al*., 2019; Zhao *et al*., 2019; Grasby *et al*., 2020). In addition, researchers begin to identify the structural and functional presentations attributed to specific genetic variants (Glahn *et al*., 2007; Hernandez *et al*., 2017; Mascheretti *et al*., 2017).

PD is associated with both genetic and environmental factors (Di Monte, 2003; Dunn *et al*., 2019; Hipp *et al*., 2019). At present, numerous genetic variants associated with PD risk have been discovered (Chang *et al*., 2017; Nalls *et al*., 2019), however, the neural mechanisms underlying PD pathogenesis of these genetic variants are largely unknown. Currently, most researchers focus on the molecular mechanisms of risk genes in PD pathogenesis using cell-based or animal-based models (Nguyen *et al*., 2011; Fusco *et al*., 2017; Burmann *et al*., 2020; Watanabe *et al*., 2020; Wie *et al*., 2021), however, whether PD-associated risk genes contribute to brain abnormalities revealed by neuroimaging studies are poorly understood. More importantly, we also don’t know whether brain network metrics mediate the associations between PD-associated risk genes and clinical manifestations of the patients. Microtube-associated protein tau (*MAPT*) rs17649553(C > T) is associated with reduced PD risk (Nalls *et al*., 2014; Georgiou *et al*., 2019) and observed to affect the expressions and splicing of multiple genes in brain tissue according to Genotype-Tissue Expression (GETx) database (https://www.gtexportal.org/home/). The goal of this study is to explore the associations between *MAPT* rs17649553 and clinical symptoms of PD patients and evaluate how *MAPT* rs17649553 affects the functional and structural networks of the patients.

Based on Parkinson’s Progression Markers Initiative (PPMI) database (www.ppmi-info.org/data), we revealed that verbal memory measured by Hopkins Verbal Learning Test– Revised (HVLT-R) was impaired in PD patients compared to control subjects as previously reported (Lucas-Jimenez *et al*., 2015; Hanoglu *et al*., 2019). Interestingly, *MAPT* rs17649553 T-carriers (CT and TT carriers) in PD patients had preserved verbal memory equivalent to that of healthy control compared to CC carriers. In this study, we determined to compare the difference of functional, white matter, and gray matter covariance networks between CC carriers and T-carriers and investigate how the functional and structural network metrics mediate the effects of *MAPT* rs17649553 on verbal memory.

## METHODS

### Study population

The raw data used in this study were downloaded from PPMI database (www.ppmi-info.org/data). The Institutional Review Board of all the participating centers approved the PPMI study and the written informed consents of participants can be obtained from the site investigators. All the research protocols of PPMI study have been published previously (Parkinson Progression Marker, 2011; Marek *et al*., 2018) and can be downloaded from the PPMI database. For up-to-date PPMI data information, visit www.ppmi-info.org. The clinical and fMRI data of PD participants were downloaded from PPMI database if the they met the inclusion criteria below: (i) The participant was diagnosed with PD according to the diagnostic criteria of International Parkinson and Movement Disorder Society; (ii) The participants acquired 3D T1-weighted MPRAGE imaging, resting-state fMRI imaging, and diffusion tensor imaging (DTI) during the same period; The participant had no other neurological diseases except PD; (iv) The participant showed no obvious structural abnormalities in both T1-weighted and T2-weighted MRI images; (v) The participant had verbal memory data assessed by HVLT-R within 3 months of fMRI imaging; (vi) The participant performed whole exome sequencing on whole-blood extracted DNA samples. According to above inclusion criteria, totally 83 PD subjects were included for the final analysis. The control subjects (n = 21) met all the inclusion criteria except the diagnosis of PD. Because most of the control subjects had no MRI data, they were not included in the neuroimaging analysis. The descriptive statistics of clinical data for PD and control subjects were shown in Table 1, Supplementary Table 1, and Supplementary Table 2.

**Table 1.** Demographic characteristics and derived scores of HVLT-R in PD patients for imaging analysis

| Characteristics | All patients<br>(n = 83) | CC carriers<br>(CC, n = 52) | T-carriers<br>(CT/TT, n = 31) | Statistic P value<br>($\chi^2$ or unpaired t-test) | Multivariate<br>regression analysis |
| --- | --- | --- | --- | --- | --- |
| Age (years) | 63.53 ± 8.24 | 63.21 ± 7.83 | 64.06 ± 8.99 | $p > 0.05$ | - |
| Sex (Female/Male) | 28/55 | 19/33 | 9/22 | $p > 0.05$ | - |
| Education (years) | 15.00 ± 2.94 | 15.21 ± 2.67 | 14.77 ± 3.10 | $p > 0.05$ | - |
| Disease duration (years) | 3.23 ± 2.57 | 3.56 ± 3.07 | 2.68 ± 1.22 | $p > 0.05$ | - |
| UPDRS-III | 22.70 ± 11.15 | 22.54 ± 11.41 | 22.97 ± 10.87 | $p > 0.05$ | $p > 0.05$ |
| MoCA | 26.41 ± 3.15 | 26.29 ± 3.27 | 26.61 ± 3.00 | $p > 0.05$ | $p > 0.05$ |
| Derived Total Recall T-score | 44.70 ± 13.77 | 42.33 ± 13.98 | 48.68 ± 12.67 | $p < 0.05^*$ | $p < 0.05^*$ |
| Derived Delayed Recall T-score | 44.64 ± 13.75 | 42.69 ± 13.98 | 47.90 ± 12.93 | $p = 0.0952$ | $p = 0.0827$ |
| Derived Retention score | 46.08 ± 13.18 | 45.71 ± 13.36 | 46.71 ± 13.08 | $p > 0.05$ | $p > 0.05$ |
| Derived Recognition Discrimination Index | 45.96 ± 12.44 | 44.98 ± 13.31 | 47.61 ± 10.82 | $p > 0.05$ | $p > 0.05$ |

\* $p < 0.05$ . Values are mean ± standard deviation. Sex ratio was evaluated with Chi-square test, other characteristics were compared using unpaired t-test. Multivariate regression analysis was performed by adjusting age, sex, disease duration, and years of education in years. *MAPT*, Microtubule-associated protein tau; HVL-T-R, Hopkins Verbal Learning Test-Revised; PD, Parkinson's disease; UPDRS-III, Unified Parkinson's Disease Rating Scales part III; MoCA, Montreal Cognitive Assessment.

### Image acquisition

The fMRI images were obtained on 3T Siemens scanners (Siemens Healthcare, Malvern, PA). The parameters for 3D T1 MPRAGE imaging were shown below: TR = 2300 ms, TE = 2.98 ms, Voxel size = 1 mm^3^, Slice thickness =1.2 mm. The T2*-weighted echo planar imaging (EPI) sequence was used for resting-state fMRI and the parameters were: TR = 2400 ms, TE = 25 ms, Voxel size = 3.3 mm^3^, Slice thickness = 3.3 mm. The parameters for DTI sequence were: TR= 8,400-8,800 ms, TE = 88 ms, Voxel size = 2 mm^3^, Slice thickness = 2 mm, 64 directions, b = 1000 s/mm^2^ along with diffusion-unweighted b0 images.

### fMRI preprocessing

Before the preprocessing of neuroimages, all the DICOM images were converted into NIFTI images using MRIcroN software (http://www.mccauslandcenter.sc.edu/mricro/mricron/). Voxel-based morphometry (VBM) was used for the processing of 3D T1 MPRAGE images, which was realized using VBM8 toolbox embedded in Statistical Parametric Mapping 12 software (SPM12, https://www.fil.ion.ucl.ac.uk/spm/software/spm12/). The 3D T1 images were normalized using Diffeomorphic Anatomic Registration Trough Exponentiated Lie algebra (DARTEL) template, and the normalized images were further segmented into gray matter, white matter, and cerebrospinal fluid (CSF). Finally, the segmented gray matter images were smoothed before they were used for the construction of gray matter covariance networks.

In order to preprocess the functional images, the first 10 volumes of images were discarded. Then, slice timing correction was performed so that all slices were temporarily aligned to a reference time point. The individual images were further realigned and registered to the first volume of images, which was spatially normalized using standard EPI template. Spatial smoothing was performed using a 4-mm Gaussian filter to improve signal-to-noise ratio and reduce anatomic variability. To reduce the effects of non-neuronal fluctuations, the head movement profile (Friston 24 parameters), CSF signals, and white matter signals were regressed out. Finally, band-pass filtering was applied to the functional image to reduce the effects of low frequency drift and high frequency physiological noise. The frequency band of time filtering was 0.01 ∼ 0.1 Hz. To adjust for the effect of head motions on functional images, we excluded subjects with head motion frame-wise displacement (FD) > 0.5 mm and head rotation > 2°. According to this criterion, nine subjects were excluded from the functional network analysis due to head movements. In addition, we also regressed the head motion parameters to adjust the effect of head motions on the functional images. We also compared the difference of head motor FD between the CC carriers and the T-carriers and found no statistical significance.

FMRIB software library toolkit (FSL, https://FSL.FMRIB.ox.ac.uk/FSL/fslwiki) was used to preprocess the DTI images. Initially, the brain skull was stripped and the brain was extracted separately using the Brain Extraction Tool (BET). In order to correct the head motion and the eddy current distortion, eddy-current correction was performed by applying a rigid-body transformation to the b0 image. Then, dtifit command of FSL was utilized to derive diffusion tensor matrix, as well as fractional anisotropy (FA), mean diffusion coefficient, axial diffusion coefficient and radial diffusion coefficient. Finally, individual images were normalized unto a standard MNI space for comparison between subjects.

### Network construction

After T1 image preprocessing, individual gray matter covariance networks were extracted using the methods proposed by Tijms et al (Tijms *et al*., 2012). This method defines network nodes as 3 × 3 × 3 voxel cubes, and network edges (referred as spatial similarity below) as correlation coefficients for gray matter morphology between each pair of nodes. We normalized gray matter covariance networks using an Automated Anatomical Labeling (AAL) parcellation atlas and created 90 × 90 gray matter covariance matrices for individuals using previously published methods (Batalle *et al*., 2013; Zhang *et al*., 2020). In order to improve the normality of the edges of gray matter covariance network, Fisher’s r-to-z transformation was applied to the correlation coefficient.

We used a free and open MATLAB toolkit, PANDA (http://www.nitrc.org/projects/panda/), to implement deterministic fiber tractography to construct the white matter network matrix. Whole-brain white matter fibers between each pair of 90 cortical and subcortical nodes in the AAL Atlas were calculated using the Fiber Assignment by Continuous Tracking (FACT) algorithm. The FA threshold was set to 0.2 and the angle threshold was set to 45°. After the tractography, a white matter network matrix based on fiber number (FN) was constructed for individual subject. We also constructed the white matter network matrix based on edge weight (EW) between two nodes of AAL atlas.

We constructed the functional connectivity matrix between individual regions in two main steps. The first step was to define 90 cortical and subcortical nodes using the AAL atlas. The second step was to calculate the pairwise function connectivity by computing the linear Pearson correlation coefficient of the time series of 90 nodes. To improve the normality of functional connectivity, we performed Fisher’s r-to-z transformation of correlation coefficients. This will create a 90 × 90 functional connectivity matrix for each subject.

### Graph-based network analysis

The topological properties of three types of networks were calculated using GRETNA (https://www.nitrc.org/projects/gretna/) (Wang *et al*., 2015). In order to compute the global and nodal network properties in each type of network, a wide range of network sparsity threshold (0.05 ∼ 0.50 with an interval of 0.05) was set. The area under curve (AUC) was calculated for each network metric which was independent of any single threshold selection. The derived global network properties included assortativity, global efficiency, local efficiency, and small-worldness metrics (clustering coefficient [Cp], characteristic path length [Lp], normalized clustering coefficient [γ], normalized characteristic path length [λ], and small worldness [σ]). The nodal network properties included nodal betweenness centrality and degree centrality. Detailed definitions of each network metric can be found in previous studies (Rubinov and Sporns, 2010; Wang *et al*., 2011).

### Statistical analysis

### Network comparison

Network-based statistic (NBS, https://www.nitrc.org/projects/nbs/) method with 5000 permutations was used to compare the global network strength of three types of networks (Zalesky *et al*., 2010). *p* < 0.05 after false discovery rate (FDR) for multiple comparisons correction was considered statistically significant (Benjamini and Yekutieli, 1995). The age, sex, years of education, and disease duration were used as covariates during the analysis.

### Comparison of network metrics

For comparison of global network metrics, two-sample permutation test (10000 permutations) was utilized. *p* < 0.05 was considered statistically significant for each sparsity threshold. For the comparison of nodal network properties and edge strengths, two-way ANOVA test was used and *p* < 0.05 after FDR correction was considered statistically significant. The AUCs of small-worldness properties were compared using unpaired t-test. Age, sex, duration of disease, and years of education were included as covariates during the statistical analysis.

### Correlation analysis

The correlation between graphic network metric and HVLT-R score was conducted by Pearson correlation method. In addition, multivariable regression analysis was used to analyze the association between graphical network metrics and HVLT-R scores by adjusting for covariates including age, sex, duration of disease, and years of education.

### Mediation analysis

The mediation analysis was performed using IBM SPSS Statistics 20. The independent variable in the mediation model was *MAPT* rs17649553 (CC, CT, and TT). The dependent variable was Derived Total Recall T-score. The mediators were graphical network metrics. We modeled the mediated relationships (indirect path) between *MAPT* rs17649553 and Derived Total Recall T-score. The model also included the direct path from *MAPT* rs17649553 to the Derived Total Recall T-score. During the mediation analysis, age, sex, disease duration, and years of education were included as covariates. *p* < 0.05 was considered statistically significant.

### eQTLs and sQTLs analysis

The data used for eQTL and sQTL analysis were downloaded from GTEx Project (https://www.gtexportal.org/home/). The normalized effect size (NES), calculated as the effect of the alternative allele (T) relative to the reference allele (C) in *MAPT* rs17649553, was obtained (https://www.gtexportal.org/home/faq). We also provided the *p*-values for the comparison of effects of two alleles on gene expression and splicing. It should be noted that only the genes reaching statistical significance in the multi-tissue comparison was shown here.

## RESULTS

### Allele and genotype frequencies

The allele frequencies (C allele frequency = 0.8072, T allele frequency = 0.1928) for *MAPT* rs17649553 were comparable to those reported in larger samples (T allele frequency = 0.2083, ALFA Project) (Molinuevo *et al*., 2016). The distribution of genotype frequencies from our participants did not deviate from Hardy-Weinberg equilibrium (*p* > 0.10). Genotype frequencies did not significantly differ with regard to age or sex (*p* > 0.05).

### Group difference of clinical variables and multivariate regression analysis

In the 83 PD participants included in this study, fifty-two subjects were CC carriers (n = 52) and the others were T-carriers (CT/TT, n = 31). The clinical characteristics of the CC carriers and T-carriers were shown in Table 1. The Derived Total Recall T-scores of HVLT-R in T-carriers were significantly higher than that of CC carriers (*p* < 0.05, Table 1). Consistently, multivariate regression analysis revealed that *MAPT* rs17649553 was significantly associated with Derived Total Recall T-scores of HVLT-R in 83 PD participants after adjusting age, sex, disease duration, and years of education. However, in control subjects, *MAPT* rs17649553 was not associated with Derived Total Recall T-scores of HVLT-R (Supplementary Table 1). The comparisons of the clinical variables including HVLT-R scores between control subjects and PD patients were shown in the Supplementary Table 2.

### Group difference of gray matter covariance network

The gray matter covariance matrices of CC carriers and T-carriers (CT and TT carriers) were shown in Fig. 1A. The NBS method showed that both positive and negative correlations of gray matter morphology significantly differ between CC carriers and T-carriers (*p* < 0.0001, NBS method, 5000 permutations, Fig. 1B). Then we used graph theory to analyze the topological metrics of gray matter covariance networks (Rubinov and Sporns, 2010; Tijms *et al*., 2012; Wang *et al*., 2015). For the global network metrics, the network assortativity was higher in T-carriers compared to CC carriers (*p* < 0.05, 10000 permutations, sparsity range: 0.30 ∼ 0.50, Fig. 1C). Both the small-worldness γ and σ were higher in CC carriers compared to T-carriers (*p* < 0.05, 10000 permutations, sparsity range: 0.20 ∼ 0.45, Fig. 1D and E). The other global network metrics were not significantly different (*p* > 0.05, data not shown). For the nodal network metrics, betweenness centrality in right medial orbitofrontal gyrus and left superior frontal gyrus were statistically different between CC carriers and T-carriers (*p* < 0.05, FDR corrected, Fig. 1F). Because betweenness centrality is a measure to evaluate the amount of influence a node has over information flow of a graph, we further assessed whether the edge strengths (spatial similarity) of right medial orbitofrontal gyrus and left superior frontal gyrus were changed by *MAPT* rs17649553. We found significantly different spatial similarity of right medial orbitofrontal gyrus and left superior frontal gyrus between T-carriers and CC carriers (*p* < 0.05, FDR corrected, Supplementary Fig. 1A-D). Additionally, we found degree centrality in multiple nodes were significantly different between CC carriers and T-carriers (*p* < 0.05, FDR corrected, Fig. 1G).

**Fig. 1.**
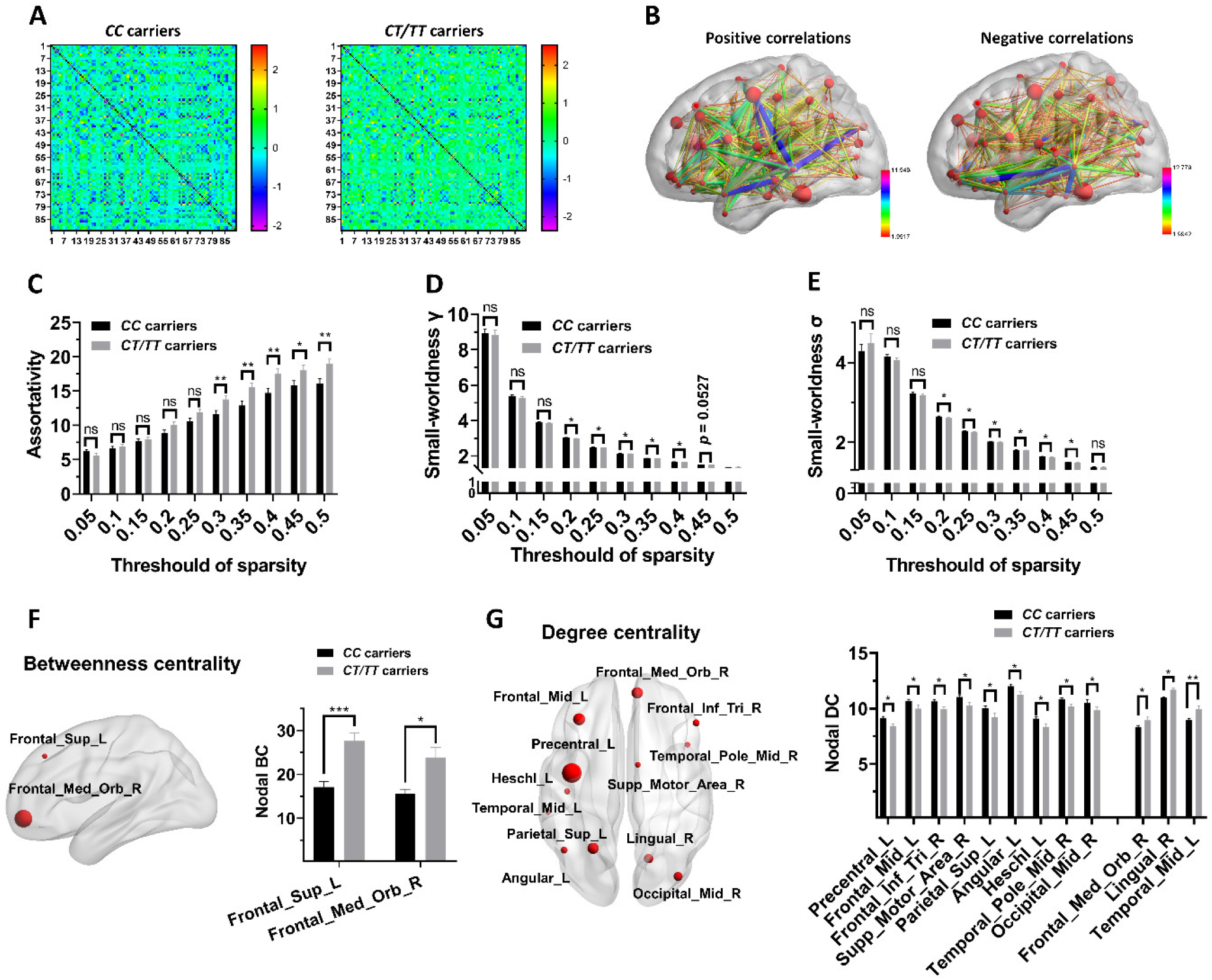
Group difference of gray matter covariance network between CC carriers and T-carriers. (A) Correlation matrix based on spatial similarity of gray matter morphology. (B) Group difference of global network spatial similarity between CC carriers and T-carriers (*p* < 0.0001, NBS method, 5000 permutations, age, sex, disease duration and years of education adjusted). (C-E) Group difference of global assortativity, small-worldness γ, and small-worldness σ (*p* < 0.05, 10000 permutations). (F) Group difference of nodal betweenness centrality between CC carriers and T-carriers (*p* < 0.05, FDR corrected). (G) Group difference of nodal degree centrality between CC carriers and T-carriers (*p* < 0.05, FDR corrected).

### Group difference of white matter network

The white matter matrices, including fiber number (FN) matrix and edge weight (EW) matrix, in CC carriers and T-carriers were shown in Fig. 2A and B. There was no statistical significance in the global connectivity strengths of FN network and EW network between CC carriers and T-carriers (*p* > 0.05, NBS method, 5000 permutations). With respect to the global topological properties of white matter network, we found the assortativity of CC carriers was significantly higher compared to T-carriers (*p* < 0.05, 10000 permutations, sparsity range: 0.15 ∼ 0.50, Fig. 2C). The small-worldness properties in EW network, including small-worldness γ, λ, and σ, were all higher in T-carriers compared to CC carriers (*p* < 0.05 or *p* < 0.01, 10000 permutations, sparsity range: 0.05 ∼ 0.50, Fig. 2D, E and F). Additionally, AUCs of small-worldness properties were also significantly different between T-carriers and CC carriers, characterized by higher AUCs of γ, λ, and σ in T-carriers compared to CC carriers (Supplementary Fig. 2A-E). For nodal network metrics, we found the betweenness centrality of left putamen was significantly higher in T-carriers compared to CC carriers in both FN network and EW network (*p* < 0.05, FDR corrected, Fig. 2G). The changes of betweenness centrality in left putamen were further demonstrated by the significant alterations of fiber amounts connecting to left putamen (*p* < 0.05, FDR corrected, Fig. 2H). We found no statistical difference in degree centrality of white matter network between CC carriers and T-carriers (*p* > 0.05, FDR corrected).

**Fig. 2.**
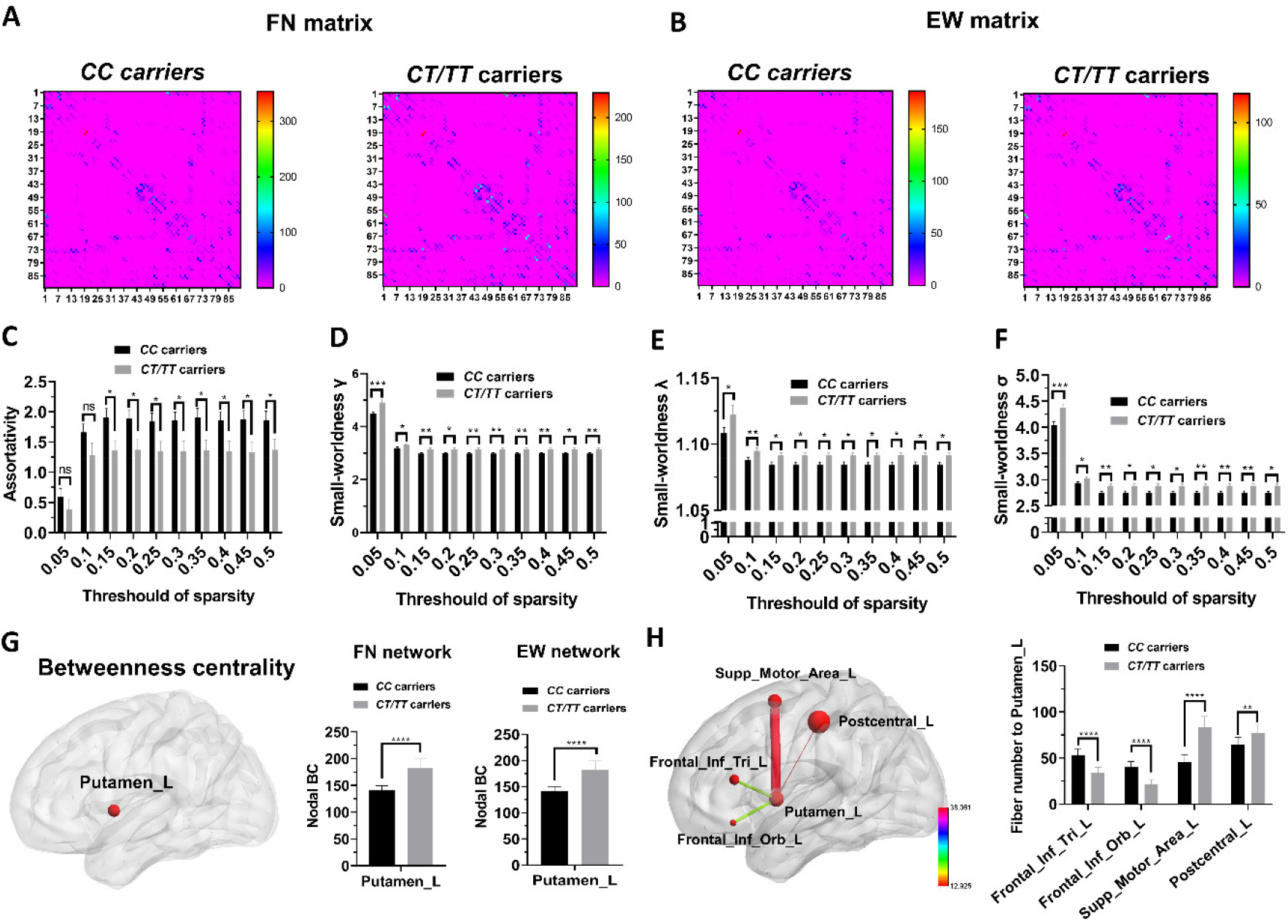
Group difference of white matter network between CC carriers and T*-*carriers. (A) Correlation matrix based on FN of white matter (*p* > 0.05, NBS method, 5000 permutations, age, sex, disease duration and years of education adjusted). (B) Correlation matrix based on EW of white matter (*p* > 0.05, NBS method, 5000 permutations, age, sex, disease duration and years of education adjusted). (C-F) Group difference of global assortativity, small-worldness γ, small-worldness λ, and small-worldness σ (*p* < 0.05, 10000 permutations). (G) Significantly different betweenness centrality in left putamen between CC carriers and T-carriers (*p* < 0.05, FDR corrected). (H) Group difference of fiber amounts connected to left putamen between CC carriers and T-carriers (*p* < 0.05, FDR corrected). FN, Fiber number; EW, Edge weight.

### Group difference of functional network

We found no significant difference on the global functional strengths of functional matrices between CC carriers and T-carriers (*p* > 0.05, NBS method, 5000 permutations, data not shown). In addition, there was no significant difference in global network properties and nodal network properties between CC carriers and T-carriers (all *p* > 0.05 in permutation test and ANOVA test, respectively, data not shown).

### Associations between gray matter covariance network metrics and verbal memory

We found the betweenness centrality of right medial orbitofrontal gyrus and left superior frontal gyrus revealed in Fig. 1F to G were not significantly correlated with derived scores of HVLT-R. However, the edge strengths between right medial orbitofrontal gyrus and left fusiform and edge strengths between left superior frontal gyrus and left precentral gyrus were correlated with Derived Total Recall T-scores (r = 0.22, *p* < 0.05 and r =0.23, *p* < 0.05, Supplementary Fig. 3A and B). In addition, we also revealed that degree centrality of right medial orbitofrontal gyrus and right supplementary motor area were correlated with derived scores of HVLT-R, as shown in Supplementary Fig. 3C-F.

### Associations between white matter network metrics and verbal memory

We found the AUC of γ, λ, and σ in both FN network and EW network were significantly correlated with Derived Total Recall T-scores (Fig. 3A-F). We also analyzed the correlations between small-worldness γ, λ, and σ spanning the sparsity of 0.05 ∼ 0.50 and Derived Total Recall T-scores of HVLT-R, we found that Derived Total Recall T-scores were also significantly correlated with small-worldness γ, λ, and σ in a wide range of sparsity threshold (Supplementary Table 3 and 4). Other network metrics of white matter network were not found to be associated with derived scores of HVLT-R.

**Fig. 3.**
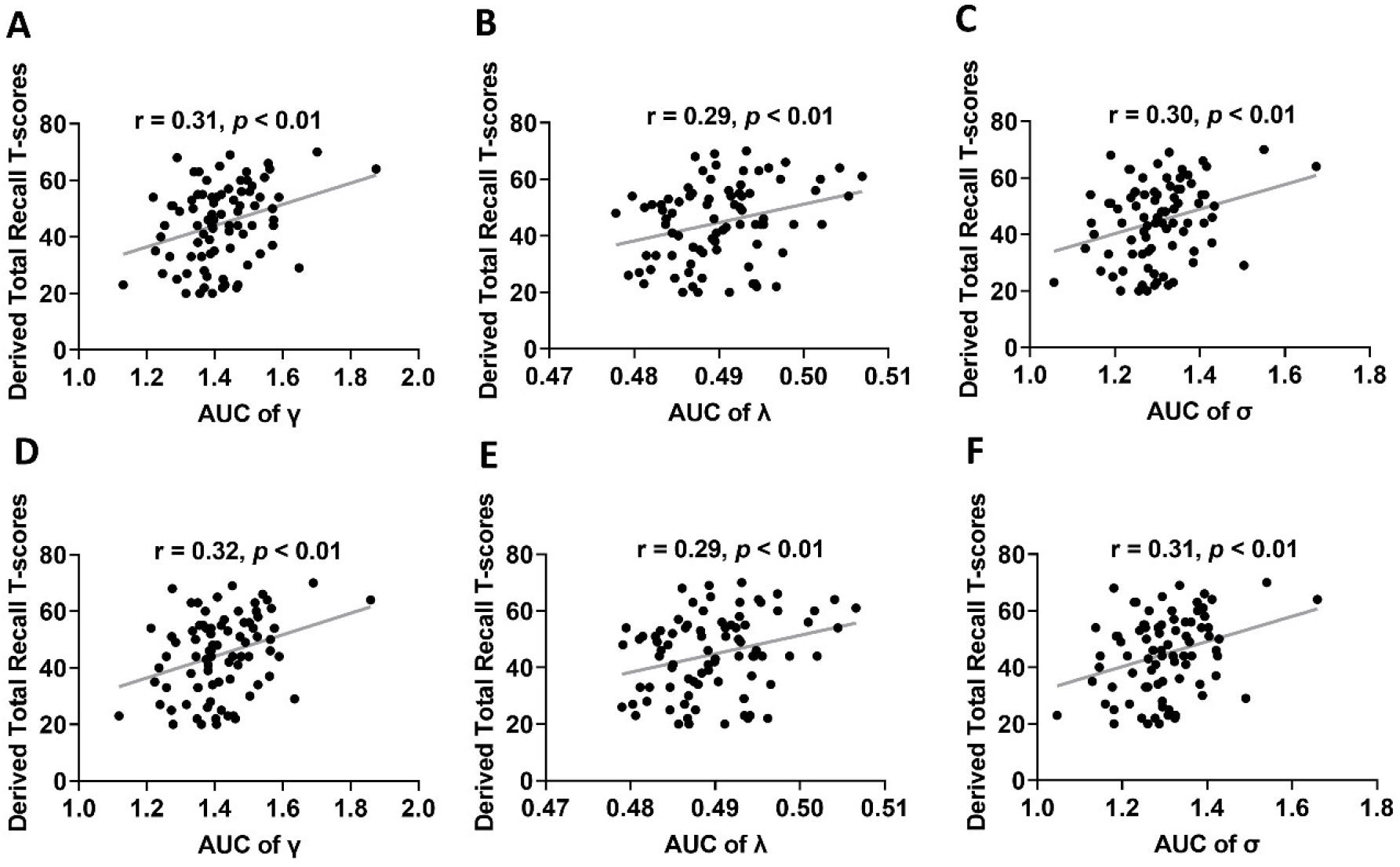
Correlations between small-worldness properties of white matter network and verbal memory. (A-C) AUCs of small-worldness γ, small-worldness λ, and small-worldness σ in FN network are correlated with Derived Total Recall T-scores. (D-F) AUCs of small-worldness γ, small-worldness λ, and small-worldness σ in EW network are correlated with Derived Total Recall T-scores. FN, Fiber number; EW, Edge weight; AUC, area under curve.

### Mediation analysis

We found AUCs of γ, λ, and σ in both FN network and EW network mediated the protective effects of *MAPT* rs17649553 on verbal memory (Fig. 4A-F). The mediation analysis of other network metrics, including gray matter covariance network metrics, all showed negative results (all *p* > 0.05).

**Fig. 4.**
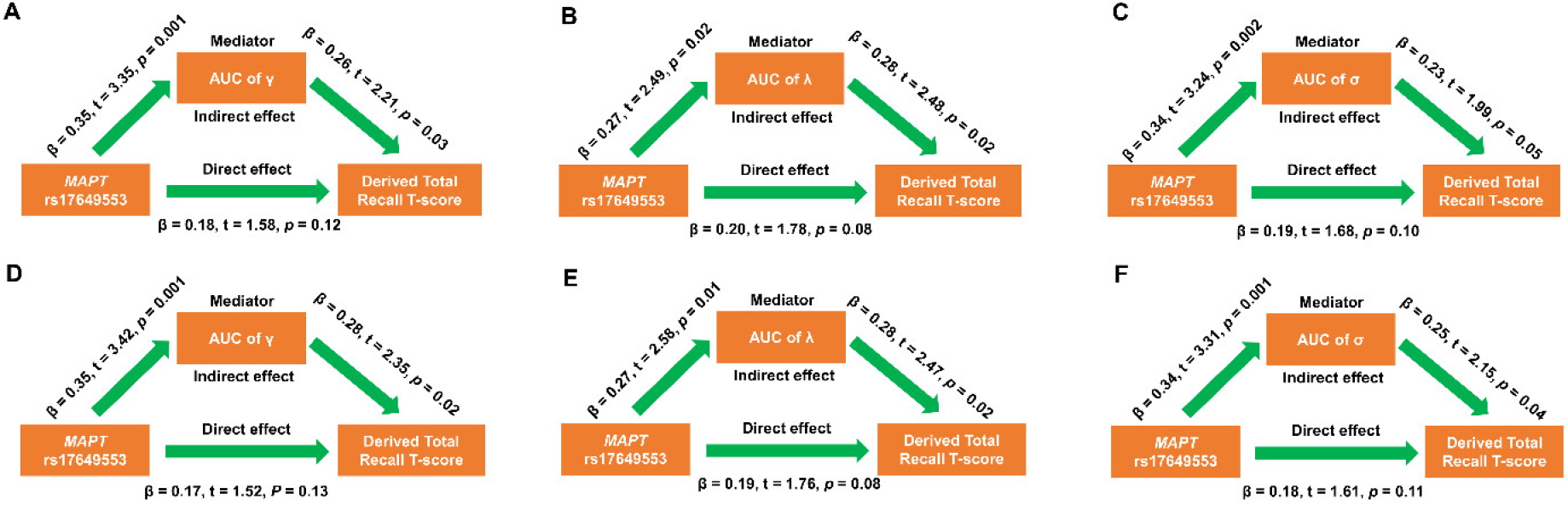
Mediation analysis suggested that small-worldness properties in white matter network mediated the protective effects of *MAPT* rs17649553 on verbal memory in PD. (A-C) Mediation analysis for AUCs of small-worldness γ, small-worldness λ, and small-worldness σ in FN network. (D-F) Mediation analysis for AUCs of small-worldness γ, small-worldness λ, and small-worldness σ in EW network. FN, Fiber number; EW, Edge weights; AUC, area under curve.

### eQTL and sQTL analysis

By analyzing the data from GTEx project, we found that *MAPT* expression was specifically increased in basal ganglia regions, including caudate (NES = 0.221, *p* = 2.2 × 10^−4^) and putamen (NES = 0.175, *p* = 0.02), but not in other brain regions (all *p* > 0.05) in T-carriers compared to CC carriers according to GTEx database.

## DISCUSSION

In this study, we identified a *MAPT* variant, rs17649553, which was significantly associated with the maintenance of verbal memory in PD, in addition to its protective effects on PD susceptibility (Nalls *et al*., 2014; Georgiou *et al*., 2019). In agreement with this finding, we found *MAPT* rs17649553 T allele modified the gray matter covariance network and white matter network towards stronger structural integrity, which may further demonstrate the preventive role of *MAPT* rs17649553 in PD risk and verbal memory impairment in PD patients. On the basis of these findings, we further showed significant correlations between structural network metrics and verbal memory. Specifically, we demonstrated that enhanced small-worldness topology of white matter network mediated the protective effects of *MAPT* rs17649553 T allele on verbal memory of PD patients.

To our knowledge, this is the first study to document that one single genetic variant can shape both the structural network topology and cognitive function, particularly verbal memory, of PD patients. Verbal memory is an important cognitive domain for humans and accumulated evidence has shown that PD patients exhibited impaired verbal memory compared to healthy control (Lucas-Jimenez *et al*., 2015; Hanoglu *et al*., 2019). Our finding of significant association between *MAPT* rs17649553 and verbal memory was novel and deserved to be further investigated in future large-scale studies. Here, we found Derived Total Recall T-scores in T-carriers were almost equivalent to that of healthy control (48.68 ± 12.67 *vs* 50.52 ± 11.85, *p* > 0.05), indicating that T allele in *MAPT* rs17649553 was a genetic predictor of verbal memory maintenance in PD patients.

We found significantly different graphical properties of gray matter covariance network between T-carriers and CC carriers, indicating that *MAPT* rs17649553 modifies the spatial topology of gray matter structure in PD patients. Interestingly, we found some of the gray matter network metrics were correlated with verbal memory as shown in Supplementary Fig. 3. These findings indicated that the covariance of gray matter morphology in right medial orbitofrontal gyrus and other brain nodes may reflect and predict the changes of verbal memory. Indeed, previous studies have implicated right medial orbitofrontal gyrus in the regulation of verbal memory in humans (Barbey *et al*., 2011; Bosch *et al*., 2013). It deserves to be noted here that unlike white matter network, the network edges in gray matter covariance network doesn’t represent real structural connectivity between nodes within this network. We consider here that the relevance of gray matter covariance network to brain behavior was still difficult to interpret and further studies were required to explore the associations between gray matter covariance network and brain behavior.

Regarding white matter network, we obtained similar profiles of global and nodal network metrics in FN network and EW network. This indicated that FN network and EW network had almost identical network topology, which was in agreement with the results of a previous study (Zhao *et al*., 2018). Our findings suggested that gray matter covariance network and white matter network exhibited divergent network topology as previously reported (Gong *et al*., 2012; Nestor *et al*., 2017). Our finding that *MAPT* rs17649553 associated with increased small-worldness γ, λ, and σ of white matter network is also novel and significant. The small-worldness properties have been found in in many biological networks, including gene transcriptional network (van Noort *et al*., 2004), protein interaction network (Bork *et al*., 2004), microRNA functional similarity networks (Le, 2015), and brain networks (Bullmore and Sporns, 2009). Interestingly, evidence has shown that small-worldness network of neurons can produce and store short-term memory (Roxin *et al*., 2004). Franzmeier et al. (2018) reported that successful memory encoding and recognition are associated with higher small-worldness of functional networks (Franzmeier *et al*., 2018). Moreover, a recent study also demonstrated increased small-worldness was associated with higher accuracy of working memory in schizophrenia (Yang *et al*., 2020). Overall, it seems that increased small-worldness corresponds to better working memory (Stevens *et al*., 2012; Langer *et al*., 2013). Furthermore, small-worldness of brain network decreased with aging (Onoda and Yamaguchi, 2013). Particularly, the reduction of small-worldness in PD patients has been reported previously (Kamagata *et al*., 2018). Therefore, we proposed that *MAPT* rs17649553 prevented verbal memory impairment in PD by increasing small-worldness of white matter network.

According to the GTEx database, *MAPT* rs17649553 was associated with the expression of both protein-encoding genes and noncoding genes, including *ARL17A, CRHR1, LINC02210, KANSL1, LRRC37A*, and *MAPT*. In fact, these genes have been associated with neuropsychiatric diseases or neurological diseases (Koolen *et al*., 2012; Zollino *et al*., 2012; Rogers *et al*., 2013; Veerappa *et al*., 2014; Jun *et al*., 2016; Meng *et al*., 2018). Particularly, the protein-coding gene *CRHR1*, has been shown to modulate anxiety behavior (Samaco *et al*., 2012), panic disorder (Weber *et al*., 2016), and depression (Dong *et al*., 2009). *KANSL1* was an essential gene involved in neurodevelopment, its mutations cause 17q21.31 microdeletion syndrome, characterized by intellectual disability and developmental delay (Koolen *et al*., 2012; Zollino *et al*., 2012). The *MAPT* gene encodes tau protein, which promotes microtubule assembly and stability, responsible for the maintenance of neuronal polarity. The variations of *MAPT* gene have been associated with AD (Desikan *et al*., 2015), PD (International Parkinson Disease Genomics *et al*., 2011), and progressive supranuclear palsy (Hoglinger *et al*., 2011). Moreover, based on GTEx database, *MAPT* rs17649553 was also linked with the splicing of *CRHR1, KANSL1, LINC02210, MAPT, LRRC37A4P, PLEKHM1*, and *ARHGAP27*. Both proteins encoded by *PLEKHM1* and *ARHGAP27* are located in the endocytosis-lysosome pathway, indicating *MAPT* rs17649553 may also affect endocytosis-lysosome functions (Tabata *et al*., 2010; Baba *et al*., 2019). Future studies were required to decode the molecular mechanisms underlying the effects of *MAPT* rs17649553 on the risk, brain networks, and verbal memory of PD.

Despite considerable strength of the analysis, limitations of our work should be acknowledged. Our study was based on the analysis of limited MRI data in single database, whether our conclusions apply to other cohort of PD patients was needed to be confirmed in future large-scale studies. Because we focus on the topological properties of structural and functional network, our methodology may be underpowered to detect focal changes of structural connectivity and functional connectivity that were also correlated with *MAPT* rs17649553. More in-depth studies, like region of interest analysis, were required to identify the focal alterations of structural network and functional network associated with *MAPT* rs17649553. Together, our results demonstrated that *MAPT* rs17649553 was associated with superior structural network topology and better verbal memory in PD.

## Data Availability

All data produced in the present study are available upon reasonable request to the authors

http://www.ppmi-info.org/data

## AUTHOR CONTRIBUTIONS

Zhichun Chen, Conceptualization, Formal analysis, Visualization, Methodology, Writing - original draft, Writing - review and editing; Bin Wu, Validation, Investigation, Methodology; Guanglu Li, Data curation, Formal analysis, Visualization; Liche Zhou, Data curation, Formal analysis, Investigation; Lina Zhang, Formal analysis, Investigation, Methodology; Jun Liu, Conceptualization, Supervision, Funding acquisition, Writing - original draft, Project administration, Writing-review and editing.

## ACKNOWLEDGMENTS

This work was supported by grants from the National Key Research and Development Program (2016YFC1306505) and the National Natural Science Foundation of China (81471287, 81071024, 81171202). Data used in the preparation of this article were obtained from the Parkinson’s Progression Markers Initiative (PPMI) database (www.ppmiinfo.org/data). We thank the share of PPMI data by all the PPMI study investigators. For up-to-date information on the study, visit www.ppmiinfo.org. PPMI – a public-private partnership – is funded by the Michael J. Fox Foundation for Parkinson’s Research and funding partners, which can be found at www.ppmiinfo.org/fundingpartners.

## DATA AVAILABILITY

All the raw data used in the preparation of this Article were downloaded from PPMI database (www.ppmi-info.org/data).All data produced in the present study are available upon reasonable request to the authors.

## SUPPLEMENTARY MATERIAL

The supplementary materials include 3 supplementary figures and 4 supplementary Tables.

