## Supplemental file for "Effects of *MAPT* rs17649553 on Structural Network Integrity and Verbal Memory in Parkinson’s Disease"

### Supplementary Material

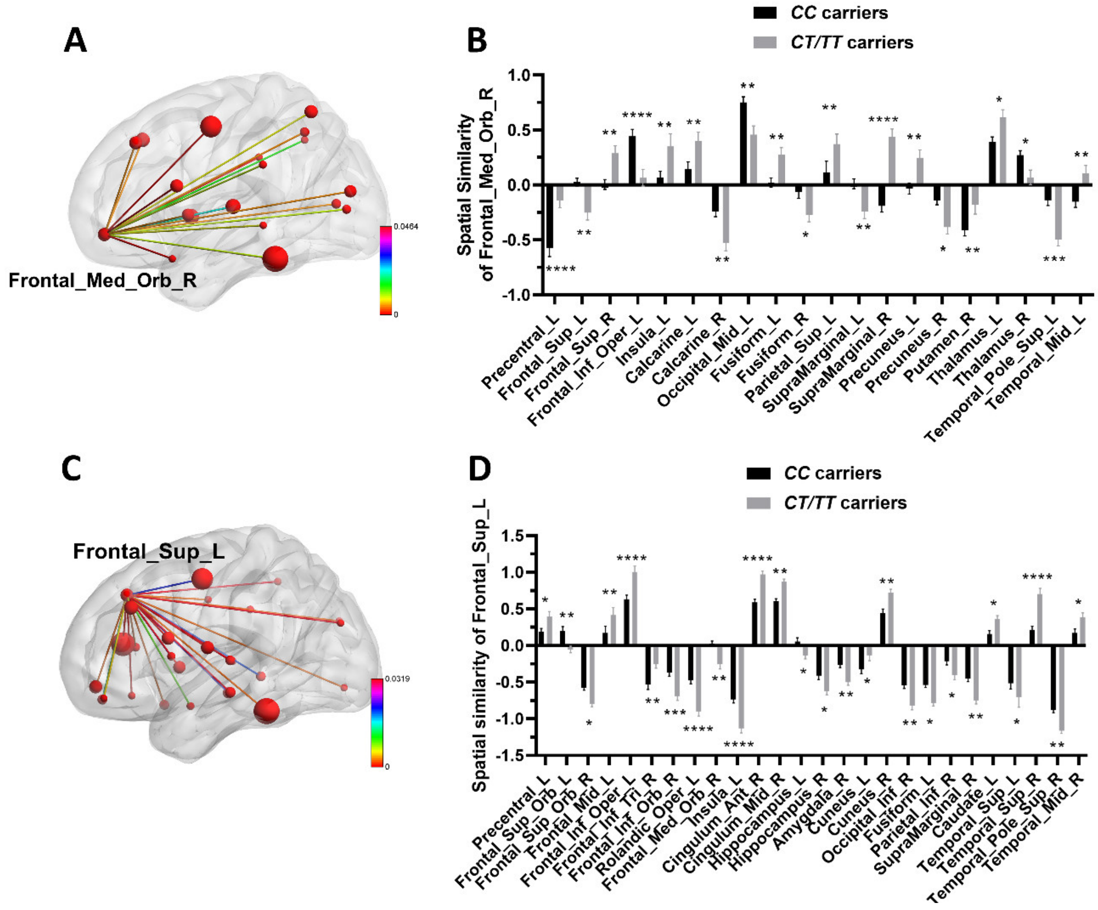


Supplementary Fig. 1. Difference in the spatial similarity of right medial orbitofrontal gyrus and left superior frontal gyrus. (A) Nodes with significantly different spatial similarity with right medial orbitofrontal gyrus between CC carriers and T-carriers. (B) Statistical difference in spatial similarity of right medial orbitofrontal gyrus (Two-way ANOVA test, FDR corrected). (C) Nodes with significantly different spatial similarity with left superior frontal gyrus between CC carriers and T-carriers. (D) Statistical difference in spatial similarity of left superior frontal gyrus (Two-way ANOVA test, FDR corrected).

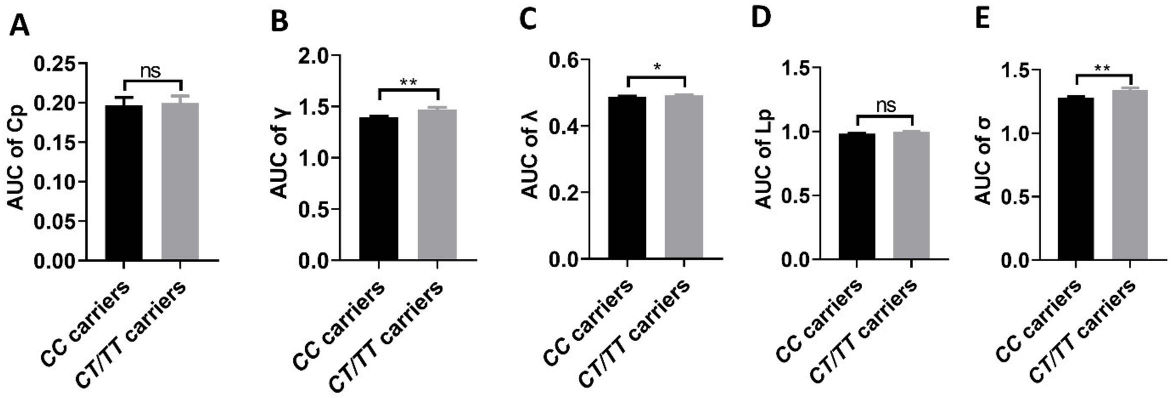


Supplementary Fig. 2. Group difference of AUCs of small-worldness properties in white matter network. (A) AUC of small-worldness Cp. (B) AUC of small-worldness γ. (C) AUC of small-worldness λ. (D) AUC of small-worldness Lp. (E) AUC of small-worldness σ. The AUCs of small-worldness properties were compared using unpaired t-test.

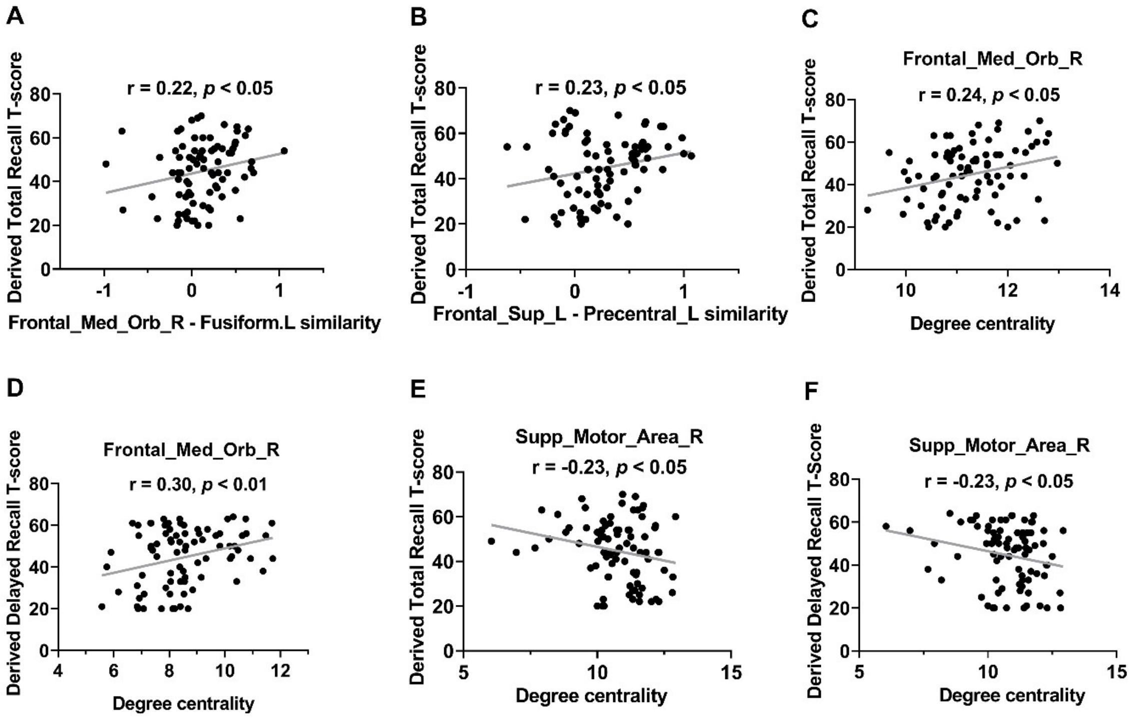


Supplementary Fig. 3. Associations between network metrics of gray matter covariance network and derived scores of HVLT-R. (A) Spatial similarity between right medial orbitofrontal gyrus and left fusiform gyrus was correlated with Derived Total Recall T-scores. (B) Spatial similarity between left superior frontal gyrus and left precentral gyrus was correlated with Derived Total Recall T-scores. (C) Degree centrality of right medial orbitofrontal gyrus was correlated with Derived Total Recall T-scores. (D) Degree centrality of right medial orbitofrontal gyrus was correlated with Derived Delayed Recall T-scores. (E) Degree centrality of right supplementary motor area was correlated with Derived Total Recall T-scores. (F) Degree centrality of right supplementary motor area was correlated with Derived Delayed Recall T-scores.

Supplementary Table 1

The correlations between verbal memory and *MAPT* rs17649553 in control subjects

| Characteristic | Control (n = 21) | Multivariate regression analysis |
|---|---|---|
| Age (years) | 62.24 ± 10.06 | - |
| Sex (Female/Male) | 5/16 | - |
| Education (years) | 16.62 ± 2.56 | $p > 0.05$ |
| MoCA | 28.14 ± 1.11 | $p > 0.05$ |
| Derived Total Recall T-score | 50.52 ± 11.85 | $p > 0.05$ |
| Derived Delayed Recall T-score | 47.29 ± 11.98 | $p > 0.05$ |
| Derived Retention score | 46.14 ± 8.95 | $p > 0.05$ |
| Derived Recognition Discrimination Index | 47.05 ± 13.07 | $p > 0.05$ |

Values are mean ± standard deviation. Multivariate regression analysis was performed by adjusting age, sex, disease duration, and years of education. *MAPT,* Microtubule-associated protein tau; MoCA, Montreal Cognitive Assessment.

Supplementary Table 2

The comparisons of clinical variables between control and PD participants

| Characteristic | Control (n = 21) | All patients (n = 83) | CC carriers (CC, n = 52) | T-carriers (CT/TT, n = 31) | Control *vs* CC carriers | Control *vs* T-carriers | Control *vs* All PD patients |
|---|---|---|---|---|---|---|---|
| Age (years) | 62.24 ± 10.06 | 63.53 ± 8.24 | 63.21 ± 7.83 | 64.06 ± 8.99 | $p > 0.05$ | $p > 0.05$ | $p > 0.05$ |
| Sex (Female/Male) | 5/16 | 28/55 | 19/33 | 9/22 | $p > 0.05$ | $p > 0.05$ | $p > 0.05$ |
| Education (years) | 16.62 ± 2.56 | 15.00 ± 2.94 | 15.21 ± 2.67 | 14.77 ± 3.10 | $p > 0.05$ | $p > 0.05$ | $p > 0.05$ |
| MoCA | 28.14 ± 1.11 | 26.41 ± 3.15 | 26.29 ± 3.27 | 26.61 ± 3.00 | $p < 0.05$* | $p > 0.05$ | $p < 0.05$* |
| Derived Total Recall T-score | 50.52 ± 11.85 | 44.70 ± 13.77 | 42.33 ± 13.98 | 48.68 ± 12.67 | $p < 0.05$* | $p > 0.05$ | $p = 0.078$ |
| Derived Delayed Recall T-score | 47.29 ± 11.98 | 44.64 ± 13.75 | 42.69 ± 13.98 | 47.90 ± 12.93 | $p > 0.05$ | $p > 0.05$ | $p > 0.05$ |
| Derived Retention score | 46.14 ± 8.95 | 46.08 ± 13.18 | 45.71 ± 13.36 | 46.71 ± 13.08 | $p > 0.05$ | $p > 0.05$ | $p > 0.05$ |
| Derived Recognition Discrimination index | 47.05 ± 13.07 | 45.96 ± 12.44 | 44.98 ± 13.31 | 47.61 ± 10.82 | $p > 0.05$ | $p > 0.05$ | $p > 0.05$ |

*$p < 0.05$. Values are mean ± standard deviation. Sex ratio was evaluated with Chi-square test, other characteristics were compared using One-way ANOVA test with Tukey's post-hoc analysis. *MAPT,* Microtubule-associated protein tau; MoCA, Montreal Cognitive Assessment; PD, Parkinson's disease.

Supplementary Table 3

The small-worldness properties of white matter FN network were correlated with verbal memory in PD patients

| Small-worldness γ | Sparsity=0.05 | Sparsity=0.1 | Sparsity=0.15 | Sparsity=0.2 | Sparsity=0.25 | Sparsity=0.3 | Sparsity=0.35 | Sparsity=0.4 | Sparsity=0.45 | Sparsity=0.5 |
|---|---|---|---|---|---|---|---|---|---|---|
| Pearson correlation | $p > 0.05$ | $p > 0.05$ | r = 0.31<br>$p < 0.01$ | r = 0.29<br>$p < 0.01$ | r = 0.31<br>$p < 0.01$ | r = 0.31<br>$p < 0.01$ | r = 0.30<br>$p < 0.01$ | r = 0.30<br>$p < 0.01$ | r = 0.31<br>$p < 0.01$ | r = 0.31<br>$p < 0.01$ |
| Multivariate linear regression | $p = 0.05$ | $p = 0.07$ | $p < 0.01$ | $p < 0.01$ | $p < 0.01$ | $p < 0.01$ | $p < 0.01$ | $p < 0.01$ | $p < 0.01$ | $p < 0.01$ |
| Small-worldness λ | Sparsity=0.05 | Sparsity=0.1 | Sparsity=0.15 | Sparsity=0.2 | Sparsity=0.25 | Sparsity=0.3 | Sparsity=0.35 | Sparsity=0.4 | Sparsity=0.45 | Sparsity=0.5 |
| Pearson correlation | $p > 0.05$ | r = 0.29<br>$p < 0.01$ | r = 0.29<br>$p < 0.01$ | r = 0.29<br>$p < 0.01$ | r = 0.29<br>$p < 0.01$ | r = 0.29<br>$p < 0.01$ | r = 0.29<br>$p < 0.01$ | r = 0.29<br>$p < 0.01$ | r = 0.29<br>$p < 0.01$ | r = 0.29<br>$p < 0.01$ |
| Multivariate linear regression | $p > 0.05$ | $p < 0.01$ | $p < 0.01$ | $p < 0.01$ | $p < 0.01$ | $p < 0.01$ | $p < 0.01$ | $p < 0.01$ | $p < 0.01$ | $p < 0.01$ |
| Small-worldness σ | Sparsity=0.05 | Sparsity=0.1 | Sparsity=0.15 | Sparsity=0.2 | Sparsity=0.25 | Sparsity=0.3 | Sparsity=0.35 | Sparsity=0.4 | Sparsity=0.45 | Sparsity=0.5 |
| Pearson correlation | $p > 0.05$ | $p > 0.05$ | r = 0.29<br>$p < 0.01$ | r = 0.28<br>$p < 0.05$ | r = 0.29<br>$p < 0.01$ | r = 0.30<br>$p < 0.01$ | r = 0.29<br>$p < 0.01$ | r = 0.28<br>$p < 0.05$ | r = 0.29<br>$p < 0.01$ | r = 0.30<br>$p < 0.01$ |
| Multivariate linear regression | $p > 0.05$ | $p > 0.05$ | $p < 0.01$ | $p < 0.05$ | $p < 0.01$ | $p < 0.01$ | $p < 0.01$ | $p < 0.05$ | $p < 0.01$ | $p < 0.01$ |

Multivariate regression analysis was performed by adjusting age, sex, disease duration, and years of education. FN, Fiber number.

Supplementary Table 4

The small-worldness properties of white matter EW network were correlated with verbal memory in PD patients

| Small-worldness $\gamma$ | Sparsity=0.05 | Sparsity=0.1 | Sparsity=0.15 | Sparsity=0.2 | Sparsity=0.25 | Sparsity=0.3 | Sparsity=0.35 | Sparsity=0.4 | Sparsity=0.45 | Sparsity=0.5 |
|---|---|---|---|---|---|---|---|---|---|---|
| Pearson correlation | $p > 0.05$ | r = 0.23<br>$p < 0.05$ | r = 0.32<br>$p < 0.01$ | r = 0.31<br>$p < 0.01$ | r = 0.30<br>$p < 0.01$ | r = 0.32<br>$p < 0.01$ | r = 0.31<br>$p < 0.01$ | r = 0.31<br>$p < 0.01$ | r = 0.32<br>$p < 0.01$ | r = 0.31<br>$p < 0.01$ |
| Multivariate linear regression | $p < 0.05$ | $p < 0.05$ | $p < 0.01$ | $p < 0.01$ | $p < 0.01$ | $p < 0.01$ | $p < 0.01$ | $p < 0.01$ | $p < 0.01$ | $p < 0.01$ |
| Small-worldness $\lambda$ | Sparsity=0.05 | Sparsity=0.1 | Sparsity=0.15 | Sparsity=0.2 | Sparsity=0.25 | Sparsity=0.3 | Sparsity=0.35 | Sparsity=0.4 | Sparsity=0.45 | Sparsity=0.5 |
| Pearson correlation | $p > 0.05$ | r = 0.29<br>$p < 0.01$ | r = 0.30<br>$p < 0.01$ | r = 0.30<br>$p < 0.01$ | r = 0.29<br>$p < 0.01$ | r = 0.29<br>$p < 0.01$ | r = 0.30<br>$p < 0.01$ | r = 0.29<br>$p < 0.01$ | r = 0.29<br>$p < 0.01$ | r = 0.29<br>$p < 0.01$ |
| Multivariate linear regression | $p < 0.05$ | $p < 0.01$ | $p < 0.01$ | $p < 0.01$ | $p < 0.01$ | $p < 0.01$ | $p < 0.01$ | $p < 0.01$ | $p < 0.01$ | $p < 0.01$ |
| Small-worldness $\sigma$ | Sparsity=0.05 | Sparsity=0.1 | Sparsity=0.15 | Sparsity=0.2 | Sparsity=0.25 | Sparsity=0.3 | Sparsity=0.35 | Sparsity=0.4 | Sparsity=0.45 | Sparsity=0.5 |
| Pearson correlation | $p > 0.05$ | $p > 0.05$ | r = 0.30<br>$p < 0.01$ | r = 0.30<br>$p < 0.01$ | r = 0.29<br>$p < 0.01$ | r = 0.30<br>$p < 0.01$ | r = 0.30<br>$p < 0.01$ | r = 0.29<br>$p < 0.01$ | r = 0.30<br>$p < 0.01$ | r = 0.30<br>$p < 0.01$ |
| Multivariate linear regression | $p < 0.05$ | $p < 0.05$ | $p < 0.01$ | $p < 0.01$ | $p < 0.05$ | $p < 0.01$ | $p < 0.01$ | $p < 0.01$ | $p < 0.01$ | $p < 0.01$ |

Multivariate regression analysis was performed by adjusting age, sex, disease duration, and years of education. EW, Edge weights.
